# Evaluating clinical utility of multi-category outcome risk prediction models

**DOI:** 10.1101/2025.04.26.25326499

**Authors:** Allison N Quintana, Christopher H Schmid, Kexin Qu, Monique Gainey, Sabiha Nasrin, Mahmuda Monjory, Eric J Nelson, Nur H Alam, Adam C Levine

## Abstract

Diagnostic models are typically evaluated by assessing their calibration and discrimination; however, neither criterion assesses the practical consequences of using a model. Decision Curve Analysis (DCA) is a method for measuring clinical utility for binary outcome models over a range of risk thresholds. While the utility of polytomous outcome models can be assessed by applying DCA to different dichotomizations of their categories, no method exists to synthesize the binary measures into a single value. This paper illustrates DCA for polytomous outcomes and extends its concepts to develop a summary utility measure for polytomous outcome models. We apply this method to three ordinal logistic regression models, including the NIRUDAK and DHAKA models for predicting dehydration severity in patients over and under five years of age, respectively.

Combining the concepts of Standardized Net Benefit (sNB) and Weighted Area Under the Net Benefit Curve, we propose the Weighted Area Under the sNB Curve (*wAUC*_*sNB*_), which can be determined for every dichotomization of a polytomous outcome. Next, we propose an average of *wAUC*_*sNB*_s weighted by the relative clinical importance of each dichotomized outcome. We term these weights importance weights and define this new measure as the Integrated Weighted Area Under the sNB Curve (*IwAUC*_*sNB*_). We apply binary DCA to the dehydration models, discuss its limitations, and apply the Integrated *wAUC*_*sNB*_ to evaluate the average utility of each model. Finally, we compare these models to criteria from the World Health Organization (WHO) and observe how the results vary for different distributional assumptions of the risk thresholds.

Applied to the NIRUDAK, DHAKA, and WHO models, the Integrated *wAUC*_*sNB*_ demonstrated that both the DHAKA and NIRUDAK models could classify individuals as benefiting from treatment better than the WHO algorithms and either of the reference strategies of treating everyone or no one.

## Introduction

Clinical prediction models that provide risk estimates for the presence of disease or a future event in the course of disease have become important tools for guiding appropriate treatment decisions. These models are typically evaluated using external validation data through measures of calibration and discrimination. Calibration refers to the agreement between the predicted and observed outcome probabilities, while discrimination refers to the model’s ability to distinguish between subjects with the outcome from those without the outcome [1]. Although necessary to the process of developing an accurate and reliable model, these measures do not assess the overall practical benefit and consequence of using the model in practice. Vickers and Elkin [2] developed Decision Curve Analysis (DCA) to assess and compare the clinical utility of prediction models by determining the relative benefits and harms associated with treatment or intervention. DCA determines the benefit of treatment given a range of reasonable risk threshold probabilities informed by clinical expertise and patient preference. The model with the largest net benefit within the range of clinically relevant thresholds has the greatest clinical utility.

Although DCA was originally developed for and has been primarily used to assess the benefit of prediction models for binary outcomes, it can be used to compare the overall utility of models for polytomous categorical outcomes that are becoming increasingly common in medical and public health applications. Existing strategies for evaluating models for nominal or ordinal outcomes decompose them into one or more binary problems [3]. One-Vs-One (OVO) approaches create decision curves comparing each pair of outcomes, whereas One-Vs-Rest (OVR) approaches compare each outcome to the others treated as a combined group. Lanzillo et al. [4], for example, assessed factors associated with both short- and long-term outcomes after rehabilitation in patients affected by acute brain injury. They used generalized ordinal logistic regression to model the short-term outcome (four classes of disability status at discharge) and performed DCA by estimating a decision curve for each of the four ordinal outcome categories versus the other three categories combined. Luo et al. [5] also developed an ordinal model to quantify the risk of obstructive sleep apnea (OSA) and constructed a single decision curve to evaluate model utility with regard to predicting any (mild, moderate or severe) versus no OSA.

The OVO and OVR approaches are primarily limited by their failure to provide a single measure that summarizes performance across all outcome categories when comparing different models. In this paper, we address this limitation by developing a new measure that summarizes the overall clinical utility of a risk prediction model for polytomous (both nominal and ordinal) outcomes. We apply the new measure and compare it to the binary approaches using ordinal diagnostic models developed to predict the amount of dehydration among patients presenting to a large urban hospital in Bangladesh [6, 7]. The resulting decision curves are then used to demonstrate the superiority of the new models to the standard World Health Organization guidelines for diagnosing the amount of dehydration.

## Materials and methods

We begin by reviewing binary and multi-category regression models with both ordered and unordered outcomes. Next, we review the concept of net benefit and decision curves for binary outcome models. Finally, we describe our extension of net benefit to polytomous outcomes and apply the method to the dehydration predictive models.

### Regression Models for Categorical Outcomes

#### Binary Logistic Regression

DCA was originally developed for a binary response variable *Y* having levels 0 and 1 for which the predicted probability of a response is described by the logistic regression model:

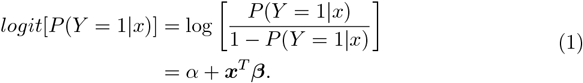

Here ***x*** is a vector of explanatory variables and ***β*** denotes a vector of the log-odds ratios associated with ***x***. Equation (1) models the log-odds of the probability that the event corresponding to outcome category 1 occurs for an individual with predictors ***x***.

### Ordinal and Nominal Logistic Regression

The cumulative logit proportional odds model

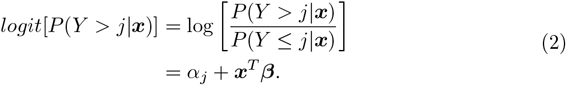

is a common one for an ordinal response *Y* with levels 0, 1, …, *K*. It models the log-odds of an observation being in a higher risk category than category *j* conditional on predictors ***x***. The set of intercepts *α*_*j*_, *j* = 0, 1, …, *K* − 1 represent the cumulative logit probabilities of an event occurring in higher outcome categories relative to lower ones when the effect of the predictors is absent. The complete model incorporates a set of *K* binary regression models in which the log-odds ratios ***β*** are the same for each binary model. In other words, the covariates ***x*** have the same effect on each set of cumulative log-odds formed as *j* changes.

When the effect of predictors may vary with the categories, the non-proportional odds model

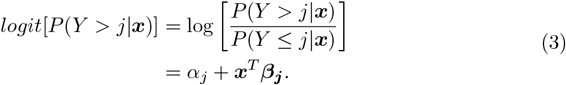

allows the regression coefficients ***β***_*j*_ to vary across the different binary splits of the response [8].

Although we focus on the cumulative logit link, all methods discussed in this paper are easily extended to other ordinal regression models with different link functions such as the cumulative probit and complementary log-log, or adjacent-category or continuation ratio formulations. [8–10]).

The methods also apply to nominal (unordered) categorical outcome models of which the most common is the baseline-category logit model [8] defined in its non-proportional odds form as:

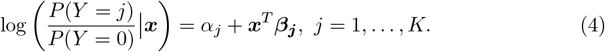

where *j* = 0 is a baseline category. The proportional odds version assumes all ***β***_***j***_ are the same. The baseline category logit model effectively subsumes *K* submodels, each comparing an outcome category to the baseline category.

### Decision Curve Analysis for Binary Outcomes

DCA depends on the concept of net benefit, which is a measure of utility [2]. We first introduce net benefit for a binary outcome.

#### Net Benefit

Suppose a clinician is uncertain as to whether or not a patient has a certain disease and, therefore, if treatment should or should not proceed. Furthermore, suppose a model has been developed to predict the probability *p* of disease for this patient. Assuming the model is correct, the patient and clinician can base a course of action for treatment upon the predicted probability. A predicted risk of disease close to one would suggest treating; a predicted risk close to zero would suggest not treating. However, the decision of whether to treat is less clear in some range of probabilities between zero and one. A decision rule or test can be formulated such that an estimated risk above some threshold probability triggers a decision to treat. Note that different patients and clinicians can have different thresholds.

Let *Y* represent true disease status (such that *Y* = 1 indicates disease-positive and *Y* = 0 disease-negative) and let *D* represent the decision to treat above some probability threshold (such that *D* = 1 indicates a decision to treat and *D* = 0 a decision not to treat). A risk probability above the threshold can then be thought of as a positive test result leading to treatment; a risk probability below the threshold is then a negative test result leading to no treatment. Each decision (test) leads to four possible results: true-positive (TP), a correct decision to treat when disease is present; true-negative (TN), a correct decision to not treat when disease is absent; false-positive (FP), an incorrect decision to treat when disease is absent; and false-negative (FN) an incorrect decision to not treat when disease is present. Each result has an associated utility denoted by *u*_*DY*_, where the first subscript represents the decision to treat (*D* = 1) or not (*D* = 0) and the second subscript indicates that disease is present (*Y* = 1) or absent (*Y* = 0). The utilities *u*_11_, *u*_10_, *u*_01_ and *u*_00_ then correspond to true-positive, false-positive, false-negative, and true-negative decisions, respectively.

To aid in understanding how these utilities reflect clinical consequences, we simply view *u*_11_ and *u*_00_ as positive because they represent utilities associated with correct decisions, and *u*_10_ and *u*_01_ as negative because they represent utilities associated with incorrect decisions. Hence the net utility of treatment for those with disease is *u*_11_ − *u*_01_; conversely, the net utility of not treating for those without disease is *u*_00_ − *u*_10_.

The expected net utility of the decision to treat is *u*_11_*p* + *u*_10_(1 − *p*): the expected utility of treatment for those with disease plus the expected utility of treatment for those without disease. Similarly, the expected net utility of the decision not to treat is *u*_01_*p* + *u*_00_(1 − *p*). At some threshold probability *p* = *r*, these expected net utilities are equal so that *u*_11_*r* + *u*_10_(− *r*) = *u*_01_*r* + *u*_00_(1− *r*) and the decision maker has equipoise about the choice to treat. Rearranging this equation gives

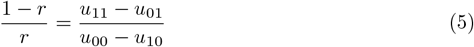

The right hand side of Equation (6) is the ratio of the net utility of treating when disease is present to the net utility of not treating when disease is absent. When Equation (6) holds under equipoise, one can determine the threshold probability *r* if one knows the net utility ratio. The ratio expresses the exchange rate a clinician places on treating diseased individuals compared with avoiding treatment of non-diseased individuals. Multiplying the numerator and denominator of the right hand side of equation (6) by −1, we also have

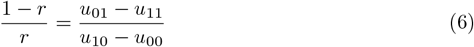

which is the net utility of not treating when disease is present to the net utility of treating when disease is absent. Intuitively, if not treating a diseased individual (a false-negative) is more dangerous than treating a non-diseased individual (a false-positive), then this ratio is greater than one and the clinician would be willing to trade off some false-positives to detect one more true-positive. Another way to think of this is as the number of non-diseased individuals the clinician would be willing to unnecessarily treat in order to treat one diseased individual. If the ratio is *K*, the clinician values a true-positive decision *K* times more than a false-positive decision and is willing to trade off *K* false-positives for every true-positive. From Equation (6), a net utility ratio of *K* corresponds to a threshold probability *r* = 1*/*(1 + *K*). Any predicted risk greater than *r* triggers a decision to treat. For example, if *K* = 9, then the threshold risk above which one would treat is 0.1.

Treatment produces a benefit when applied to diseased individuals, the true-positives, and produces harm when applied to non-diseased individuals, the false-positives. The Net Benefit (NB) of treatment is the weighted difference between the proportion of true-positives (*TP/N*) and false-positives (*FP/N*) in the population of interest of size *N*, with weights corresponding to the relative utility of treating diseased and non-diseased individuals. Setting the weight for *TP/N* as one, the weight for *FP/N* is the relative utility of treating non-diseased compared to diseased individuals (i.e., the inverse of the relative utility of treating diseased compared to non-diseased individuals). The NB of treatment for a specific threshold *r* may be written as

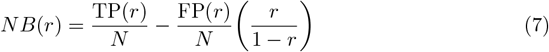

where *TP* (*r*) and *FP* (*r*) are the true- and false-positive counts, respectively, when the test result is defined as positive for risks above the threshold *r*. Equation (7) can also be interpreted in terms of costs and benefits with *r/*(1 − *r*) representing the ratio of costs to benefits with the assumption that each *TP* has the same expected benefit and each *FP* has the same expected cost independent of the predicted risk [11].

The equation may be rewritten in terms of model sensitivity *se*(*r*), specificity *sp*(*r*), and disease prevalence *P* [3] as

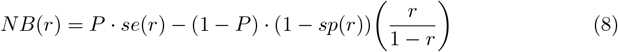

where,

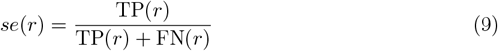

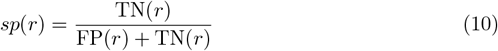

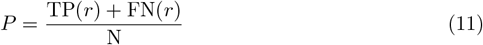

and *FN* (*r*) and *TN* (*r*) are the corresponding false- and true-negative counts, respectively, for a threshold of *r*. For a fixed risk threshold *r*, one can determine the NB provided by a model or some other treatment-guiding strategy. Note that the units of NB are true-positives. Therefore, a NB of 0.05 implies that the model identifies five more true-positive results than discounted false-positive results for every 100 individuals in the target population.

We can determine the NB using the estimated probabilities from a prediction model or from the presence of a disease biomarker.

Alternatively, the NB can also be determined for the clinical strategies of treating all individuals and treating none. “Treat All” and “Treat None” are reference strategies included in every DCA. They represent scenarios that assume either all individuals are diseased so that everyone should be treated, or that no individuals are diseased so that none should be treated. DCA compares the NB of a prediction model (or biomarker) to that of both reference strategies because Treat All and Treat None may be reasonable clinical strategies if the risk threshold is either very low or very high, respectively.

The strategies of Treat All and Treat None are quantified based on the NB implicitly associated with their assumptions. By definition, Treat None has zero true-positive and false-positive counts. Sensitivity is therefore zero and specificity is one. Because no individuals are treated, treatment gives no benefits or costs, and hence *NB*_Treat None_ = 0 for all thresholds *r*. In essence, the Treat None strategy makes sense only for the clinician whose threshold to treat is set at *r* = 1. In contrast, the Treat All strategy has sensitivity one and specificity zero. The true-positive and false-positive counts are simply the number of individuals with and without disease, respectively. The proportion of true-positives is simply the prevalence, *P*, and the proportion of false-positives is 1 − *P*. Therefore,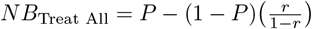. This is optimized at *P* when *r* = 0.

### Decision Curves for Binary Outcomes

Decision curves evaluate the NB of using a prediction model to guide treatment decisions over a range of thresholds representing the relative harms of false-positive and false-negative decisions. They are constructed by plotting NB against a range of risk threshold probabilities *r* or odds (cost-benefit ratios) *r/*(1− *r*). Typically, a decision curve should only be evaluated over risk thresholds of clinical relevance that reflect a decision maker’s propensity for choosing treatment. For example, a clinician weighing the risks and benefits of biopsy for detecting cancer may be willing to perform up to 10 biopsies to detect one cancer [12]. Thus, if a prediction model estimates a patient’s risk for cancer to be 10% or greater, the clinician would biopsy; otherwise, she would not biopsy and would instead just monitor the patient’s health. Other clinicians may have different risk thresholds so it is useful to show the curve over a region of thresholds that reasonable clinicians might have in a specific clinical context.

Decision curves should be read vertically at a particular risk threshold because net benefit quantifies the benefit accruing given a specific cost-benefit ratio. If we compare two points on a curve at different thresholds, then we are comparing the net benefit for individuals with different cost-benefit ratios. If we compare the net benefit for different curves (corresponding to two different risk prediction models), then we must fix the cost-benefit ratio to have a fair comparison [11].

A test or model that offers value in clinical practice should have a positive NB. Thus, when comparing decision curves from two different models, the model with higher NB in the range of reasonable thresholds provides greater utility. For a fixed risk threshold, greater NB signifies that the net increase in the proportion of appropriately treated patients is greater for a model with larger NB than for a model with smaller NB [13].

While a decision curve displays the net benefit of a decision model over a set of relevant clinical thresholds, we are faced with several potential problems when we wish to compare the net benefit of two or more models expressed as decision curves across a range of clinically relevant thresholds. First, the decision curves may not be well-ordered. In other words, the curves might cross indicating that one model is preferred for certain thresholds and another for different ones. While each clinician with a specific threshold can choose one optimal model, determining the best model for a group of clinicians with different thresholds is not so straightforward. This becomes even more complicated for multiple or polytomous outcomes when different models may dominate across the relevant threshold range for different sets of outcomes or outcome categories.

To compare different models, we clearly need some measure of total net benefit calculated across the range of relevant clinical thresholds. Such a measure must also account for the relative importance attached by different clinicians to potential relevant threshold probabilities.

### Area Under the Net Benefit Curve

A natural measure of total net benefit that addresses the problem of comparing two models in which neither dominates the other over the entire range of risk thresholds is the Area Under the Net Benefit Curve

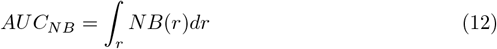

proposed by Talluri and Shete [14] to summarize the performance of a model over the clinically-chosen range of risk threshold probabilities. It is analogous to the area under a receiver operating characteristic (ROC) curve commonly used to represent discrimination between different outcome categories in binary regression.

The central limitation of the *AUC*_*NB*_ is its assumption of a uniform distribution of threshold probabilities over which to integrate NB. In other words, *AUC*_*NB*_ assumes that a set of decision makers choose their thresholds uniformly over a given range. Another interpretation is that a single decision-maker weights all thresholds equally. In reality, a single clinician certainly prefers certain thresholds over others (and in fact may have only one threshold) and a set of clinicians prefers thresholds in the middle of the range compared to the extremes. For this reason, Talluri and Shete [14] extended *AUC*_*NB*_ by weighting the area by the distribution *f* (*r*) of preferences over clinically-relevant risk thresholds to form the Weighted Area Under the NB Curve

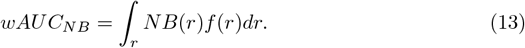

The maximum achievable NB is the outcome prevalence, achieved by using a model that perfectly discriminates those who have disease from those who do not and has both sensitivity and specificity equal to one. This limiting value complicates the comparison of decision curves for outcomes or populations with different prevalences. Standardizing net benefit by prevalence to form standardized NB *sNB* ≡ *NB/P* [11] (also known as relative utility [15, 16]) solves this problem. The maximum value of sNB is 1; thus, sNB can be interpreted as the proportion of the maximum NB achieved. We can then rewrite equation (8) as

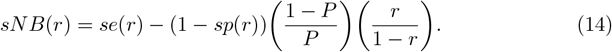

Combining sNB with weighting by *f* (*r*), we finally arrive at the Weighted Area Under the Standardized Net Benefit Curve

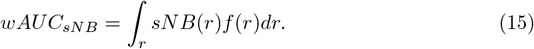

### Decision Curve Analysis for Polytomous Outcomes

We now extend net benefit to polytomous outcomes **Y** = (*Y*_0_, …, *Y*_*K*_) with *K* + 1 categories. Let **p** be the *N ×* (*K* + 1) matrix of outcome probabilities for **Y** having elements *p*_*ij*_ representing the probability of outcome category *j* = 0, 1, …, *K* for individual *i* = 1, 2, …, *N*.

The fundamental connection between decision curves for binary and polytomous outcomes follows from noting that models for an outcome with *K* categories can be written as *K* − 1 component binary comparisons. For cumulative logit ordinal regression models with three outcome categories, the binary comparisons are *p*_*i*0_ vs. *p*_*i*1_ + *p*_*i*2_ and *p*_*i*0_ + *p*_*i*1_ vs. *p*_*i*2_ which are of an OVR form. With *K >* 3 categories, however, only the first and last comparisons, *p*_*i*0_ vs. *p*_*i*1_ + … + *p*_*iK*_ and *p*_*i*0_ + … + *p*_*i,K*−1_ vs. *p*_*iK*_, are OVR. The remaining *K* − 2 comparisons *p*_*i*0_ + *p*_*i*1_ + … + *p*_*ik*_ vs. *p*_*i,k*+1_ + … + *p*_*iK*_ for *k* = 1, …, *K* − 2 are of a Some Versus Rest (SVR) form. In a baseline-category logit regression model, all comparisons to the baseline category 0 take an OVO form *p*_*ij*_ vs. *p*_*i*0_ for *j* = 1, …, *K*.

Each component binary comparison leads to a single binary decision curve. Let 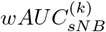 be the weighted area under the standardized net benefit curve for the *k*^*th*^ component binary model. This can be interpreted as the average benefit attached to this outcome comparison across a group of clinicians with different risk thresholds or as the benefit for a single clinician averaged over the clinician’s risk threshold preference distribution. DCA can, therefore, be applied to polytomous responses by applying binary DCA to these sets of binary models [3]. Both OVR and OVO techniques have been employed in practice; however, OVR is more common.

The difficulty with using DCA separately for each binary comparison, however, is that one cannot obtain a single decision curve for the polytomous outcome. To properly combine the separate binary decision curves into a single measure that describes the net benefit for a model of a polytomous outcome, we need to average net benefit across the different binary models while properly accounting for the potentially different clinical importance of each outcome. For instance, preventing a severe outcome is more important than preventing a less severe outcome and most clinicians would choose a much lower threshold at which to treat the severe outcome. Therefore, it seems reasonable to weight the areas under the different curves by the relative importance of the outcomes to different clinicians.

We refer to these weights as importance weights and denote them by *w* = (*w*_1_, …, *w*_*K*_), standardizing them so that *w*_*k*_ ∈ [0, 1] and 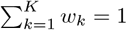. No weight is needed for the reference category. As with the risk thresholds *r*_*k*_, different clinicians can have different importance weights and single clinicians might wish to represent uncertainty about their own weights. This uncertainty can be represented by a probability distribution of importance weights, *f* (*w*).

Consider first the integrated standardized net benefit for an individual clinician operating at a risk threshold *r*_*i*_ with importance weights *w*_*i*_ = (*w*_*i*1_, …, *w*_*iK*_)

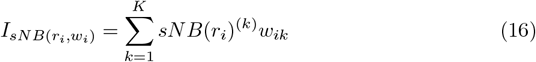

where *sNB*(*r*_*i*_)^(*k*)^ are the standardized net benefits for *K* binary comparisons.

To obtain the total net benefit across a group of clinicians, we need to average these individual integrated benefits across the distributions of clinician thresholds *f* (*r*) and clinician importance weights *f* (*w*). This gives the Integrated Weighted Area Under Standardized Net Benefit Curves

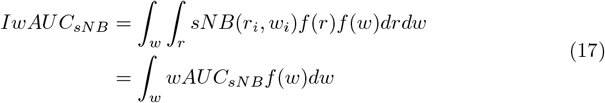

The measure can be applied to any polytomous outcome regression model, whether the outcome is ordered or unordered and with any link function. Given that weights are being applied to categorical events, a reasonable distributional form for *f* (*w*) might be a beta distribution for *K* = 2 or a Dirichlet distribution when *K >* 2.

The measures represented by all of these integrals can be computed with Monte Carlo integration; standard errors and confidence intervals for the empirical estimates can be computed using bootstrap resampling [17].

### Dehydration Study Data

To illustrate the methods presented in this paper, we use ordinal regression models fit to data from two studies conducted in Bangladesh to predict dehydration severity in two separate cohorts. The Dehydration: Assessing Kids Accurately (DHAKA) and Novel, Innovative Research for Understanding Dehydration in Adults and Kids (NIRUDAK, meaning “dehydrated” in Bangla) studies were prospective cohort studies of patients under and over five years of age, respectively, presenting with acute diarrhea to the International Centre for Diarrhoeal Disease Research, Bangladesh (icddr,b) Dhaka Hospital. The studies aimed to create clinical diagnostic models for assessing dehydration in a resource-limited setting [6]. Study enrollment took place in the icddr,b rehydration unit between February and June 2014 for the DHAKA cohort and between March 2019 and March 2020 for the NIRUDAK cohort. Ethical approval for both the DHAKA Study and the NIRUDAK study were obtained from the icddr,b’s Research Review Committee and Ethical Review Committee and the Rhode Island Hospital’s Institutional Review Board. All methods were performed in accordance with relevant guidelines and regulations. Formal written consent was obtained from each participant and/or their parent/guardian if under 18 years old in their native language, Bangla. Baseline data collection included weight measurement, assessment of ten clinical signs of dehydration (general appearance, skin pinch, sunken eyes, tears, radial pulse, deep breathing, extremity warmth, heart rate, mucous membranes, capillary refill), as well as demographic information. Individuals in the DHAKA and NIRUDAK cohorts were weighed every eight and four hours, respectively, to determine their post-hydration stable weight. This stable weight was determined by averaging the two highest consecutive weight measurements that differed by less than 2%. The outcome was percent dehydration, calculated as percent weight change after rehydration for individuals with a valid stable weight as

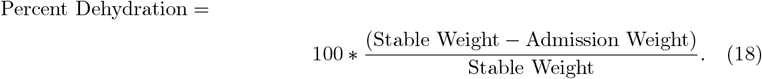

Three ordinal dehydration categories were formed by categorizing percent dehydration as severe (*>* 9%), some (3-9%), and no (*<* 3%) dehydration.

Levine et al. [6, 7] used cumulative logit proportional odds ordinal logistic regression to model this categorical outcome for children under five years in the DHAKA model [6] (Table 1) and for adults and children over five years in two different NIRUDAK models, a full model and a simplified model [7] (Table 2).

**Table 1.** DHAKA model.

| Clinical Sign | Regression Coefficients | OR (95% CI) |
| --- | --- | --- |
| **General Appearance** |  |  |
| Normal (reference) | 1 | 1 |
| Restless/Irritable | 0.72 | 2.06 (1.42, 2.99) |
| Lethargic/Unconscious | 1.23 | 3.41 (2.24, 5.23) |
| **Respiration Depth** |  |  |
| Normal (reference) | 1 | 1 |
| Deep | 0.47 | 1.60 (1.08, 2.35) |
| **Skin Pinch** |  |  |
| Normal (reference) | 1 | 1 |
| Slow | 0.82 | 2.27 (1.61, 3.22) |
| Very Slow | 1.37 | 3.94 (2.10, 7.41) |
| **Tears** |  |  |
| Normal | 1 | 1 |
| Decreased | 0.34 | 1.41 (1.02, 1.94) |
| Absent | 0.74 | 2.09 (1.21, 3.61) |

**Table 2.** NIRUDAK models.

| Clinical Sign | Regression Coefficients | OR (95% CI) |
| --- | --- | --- |
| **Full Model** |  |  |
| Age (per 1 yr) | -0.01 | 0.99 (0.99, 1.00) |
| **Skin Pinch** |  |  |
| Rapid (reference) | 1 | 1 |
| Slow | 0.71 | 2.03 (1.60, 2.58) |
| Very Slow | 1.53 | 4.60 (3.15, 6.54) |
| **Eye Level** |  |  |
| Normal (reference) | 1 | 1 |
| Sunken | 0.70 | 2.02 (1.61, 2.55) |
| **Respiration Depth** |  |  |
| Normal (reference) | 1 | 1 |
| Deep | 0.37 | 1.45 (1.17, 1.79) |
| Vomiting Episodes in 24hr. |  |  |
| <1 (reference) | 1 | 1 |
| 1-5 | 0.39 | 1.48 (1.07, 2.07) |
| 6-10 | 0.69 | 1.99 (1.42, 2.79) |
| >10 | 0.91 | 2.48 (1.71, 3.60) |
| **Systolic BP Flat**<br>(per 10mmHg increase) | -0.14 | 0.87 (0.82, 0.91) |
| **MUAC**<br>(per 10mm increase) | -0.09 | 0.92 (0.89, 0.94) |
| **Sex** |  |  |
| Female (reference) | 1 | 1 |
| Male | 0.36 | 1.43 (1.19, 1.73) |
| **Intercept** |  |  |
| Any Dehydration | -3.38 | 0.03 |
| Severe Dehydration | 0.66 | 1.94 |
| **Simplified Model** |  |  |
| **Skin Pinch** |  |  |
| Rapid (reference) | 1 | 1 |
| Slow | 0.68 | 1.98 (1.58, 2.49) |
| Very Slow | 1.34 | 3.83 (2.72, 5.40) |
| **Eye Level** |  |  |
| Normal (reference) | 1 | 1 |
| Sunken | 0.60 | 1.82 (1.46, 2.29) |
| **Respiration Depth** |  |  |
| Normal (reference) | 1 | 1 |
| Deep | 0.58 | 1.79 (1.45, 2.20) |
| **Urine Output** |  |  |
| Normal (reference) | 1 | 1 |
| Decreased | 0.29 | 1.33 (0.97, 1.82) |
| Minimal | 0.58 | 1.78 (1.18, 2.69) |
| **Radial Pulse** |  |  |
| Strong (reference) | 1 | 1 |
| Decreased | 0.44 | 1.55 (1.25, 1.93) |
| Absent | 1.07 | 2.91 (1.53, 5.49) |
| **Intercept** |  |  |
| Any Dehydration | 0.29 | 1.34 |
| Severe Dehydration | 4.08 | 59.09 |

#### World Health Organization Guidelines

The World Health Organization (WHO) has developed an algorithm referred to as the WHO Integrated Management of Childhood Illness (IMCI) for determining dehydration severity in children under five years with acute diarrhea [18]. The WHO slightly modified the IMCI algorithm for adults and older children to create a simple algorithm called the WHO Integrated Management of Adolescent and Adult Illness (IMAI) [19]. These algorithms use the same clinical symptoms as assessed in the DHAKA and NIRUDAK studies to classify individuals in one of the three ordinal dehydration categories (see Table 3). As these algorithms are widely used for determining dehydration severity, we compare their clinical utility to that of the models associated with the NIRUDAK and DHAKA studies.

**Table 3.** WHO IMAI algorithm for dehydration assessment [7, 19].

| Signs | Classify as | Treatments |
| --- | --- | --- |
| Two of the following signs:<br>• Lethargic or unconscious<br>• Sunken eyes<br>• Not able to drink or is drinking poorly<br>• Skin pinch goes back very slowly | SEVERE DEHYDRATION | Rehydration with IV or NG – Plan C<br>Consider causes and treat. If there is cholera in your area, give appropriate antibiotic for cholera (according to sensitive data).<br>Report cases (see Section 21 Recording and reporting disease outbreaks). |
| Two of the following signs:<br>• Sunken eyes<br>• Drinks eagerly, thirsty<br>• Skin pinch goes back slowly | SOME DEHYDRATION | Give fluid and food – Plan B<br>Immediately advise patient when to return. Follow up in 5 days if not improving. |
| Not enough signs to classify as severe or some dehydration. | NO DEHYDRATION | Treat diarrhoea at home – Plan A<br>Advise when to return.<br>Follow up in 5 days if not improving. |

## Results

We illustrate the use of these methods by first applying binary DCA to dichotomizations of the ordinal outcomes for the DHAKA and NIRUDAK data, comparing the new models to the WHO algorithms. Then we implement the new measure *IwAUC*_*sNB*_ to compare the models across all three outcome categories.

Consider the binary outcome of severe dehydration. Clinically speaking, determining whether an individual is severely dehydrated is critical because it requires admission to a hospital for treatment with intravenous fluids. Thus, it is reasonable to examine this category on its own. Collapsing the categories of some and no dehydration into a single category, we compare severe versus not severe (some or no) dehydration. For each individual, we sum their predicted probabilities of some and no dehydration from the ordinal model to form a single predicted probability for the collapsed category.

A clinician may also be interested in distinguishing between any (severe or some) and no dehydration, particularly in an environment with limited resources where over-treatment may also incur high societal costs [7]. Consequently, “any dehydration” appears to be a reasonable outcome of interest. Thus, we also collapse the ordinal categories of some and severe dehydration by summing the ordinal predicted probabilities to obtain a binary comparison of any versus no dehydration.

In the context of OVR DCA, one might be tempted to also consider other binary comparisons, such as “some dehydration” as an outcome of interest by collapsing severe and no dehydration into a single category. However, this choice is only meaningful in practice for unordered outcomes; in ordinal regression models, only adjacent or cumulative categories are compared with one another.

Based on the predicted probabilities from the two binary regression models of severe and any dehydration, we can construct two sets of decision curves for the DHAKA, NIRUDAK and WHO models. Fig 1 gives the decision curves for the DHAKA and WHO IMCI models as well as the Treat All and Treat None strategies for both severe and any dehydration for children under 5. Fig 2 gives the corresponding curves for the adult NIRUDAK and WHO IMAI models. Subfigures (a) and (c) in each figure display the curves for the entire range of possible thresholds and subfigures (b) and (d) show them only over ranges that a clinician might believe to be relevant for each outcome: 0.05 to 0.15 (5 to 15%) for severe dehydration, and 0.6 to 0.8 (60 to 80%) for any dehydration.

**Fig 1.**
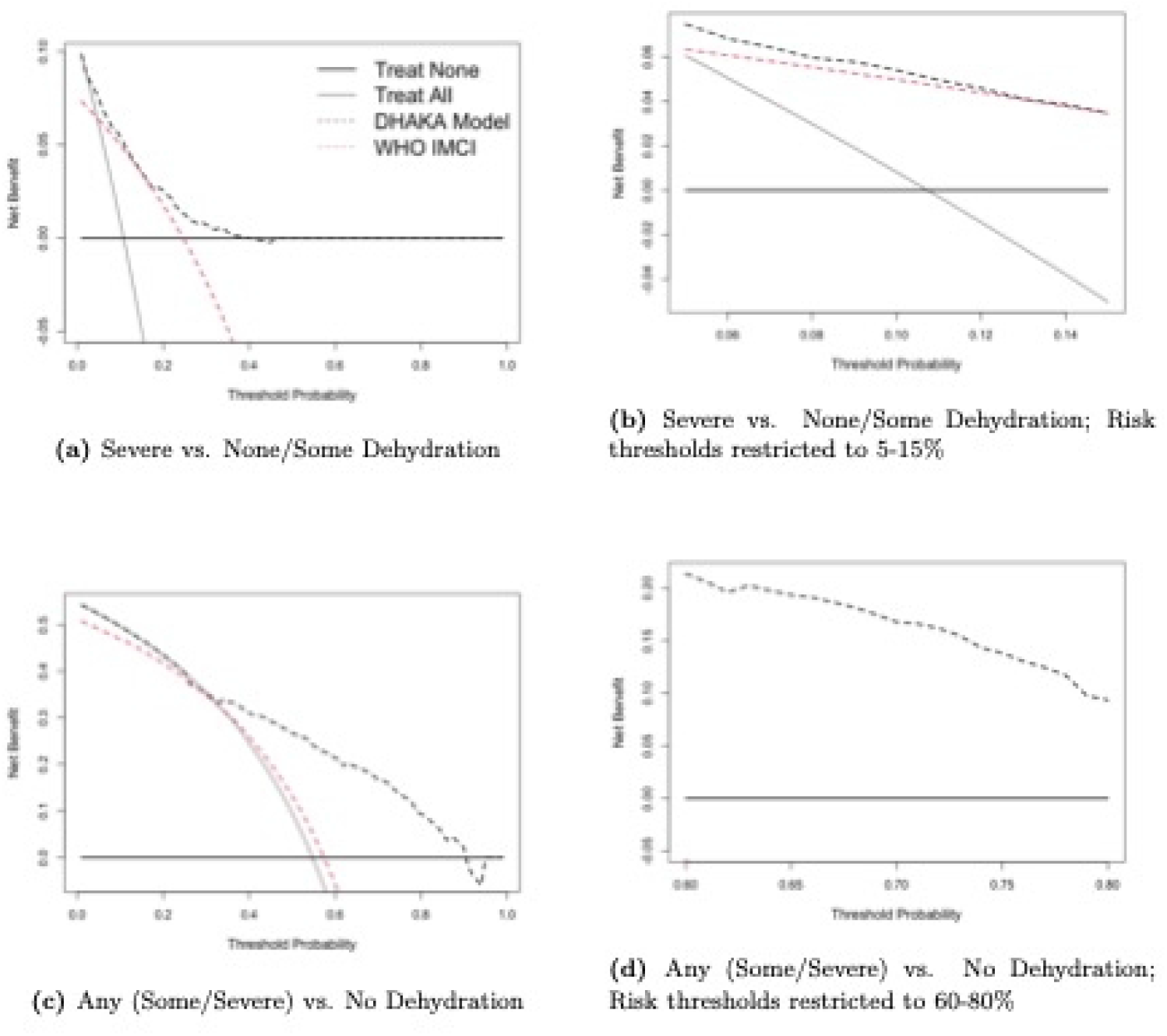
Decision curve analysis for DHAKA ordinal model and WHO IMCI algorithm (for individuals ages *<* 5 years).

**Fig 2.**
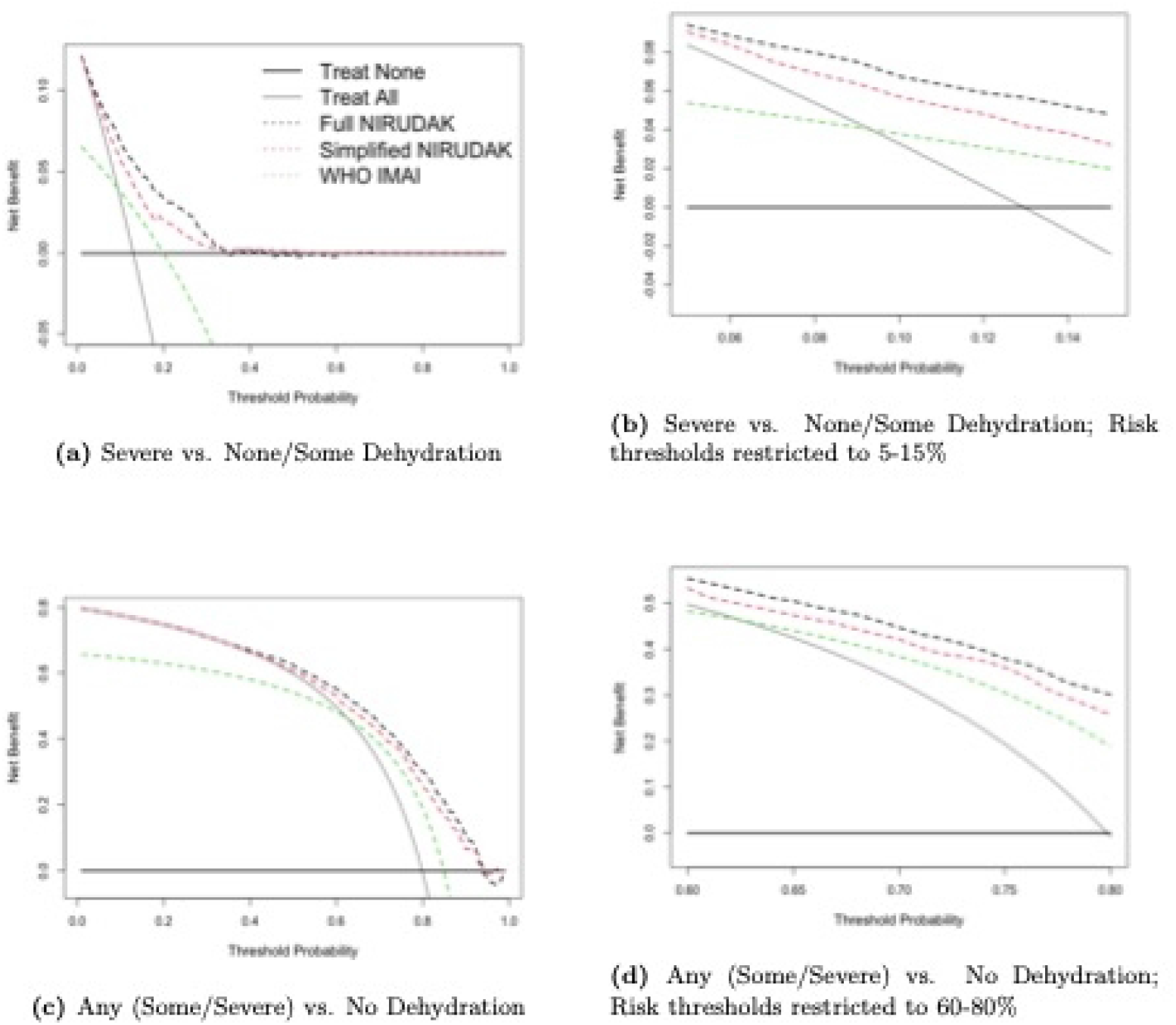
Decision curve analysis for NIRUDAK ordinal models and WHO IMAI algorithm (for individuals ages *>* 5 years).

These ranges reflect the relative odds clinicians assign to missing a case of severe (or any) dehydration compared with providing unnecessary resuscitation with intravenous fluids (or treating a patient unnecessarily). A range of 0.05 to 0.15 corresponds to odds of 5.7 to 19 in favor of treating, indicating the large relative harm of missing a severe case. Conversely, a range of 0.6 to 0.8 corresponds to odds of 1.5 to 4 in favor of not treating, indicating the much lower relative harm of not treating a less severe case. The appropriate range of risk thresholds is lower for more severe outcomes and higher for less severe outcomes, reflecting the tendency for greater risk-aversion with more severe illness.

Sub-figures (b) and (d) in Fig 1 and Fig 2 show that the DHAKA and NIRUDAK models have greater net benefit than the WHO IMCI and IMAI algorithms as well as the Treat All and Treat None strategies across the range of appropriate thresholds for severe versus non-severe dehydration and for any versus no dehydration. In fact, the DHAKA model is so much better that the decision curves for the WHO IMCI algorithm and Treat All strategy are not even visible in Fig 1(d) as both have a negative NB in this range.

Fig 3 shows the standardized net benefit curves, repeating Figs 1(a) and 1(c) as well as Figs 2(a) and 2(c) but with the y-axis scaled by the prevalence. The maximum value is now 1, although the curves can still have negative values when the net benefit is negative.

**Fig 3.**
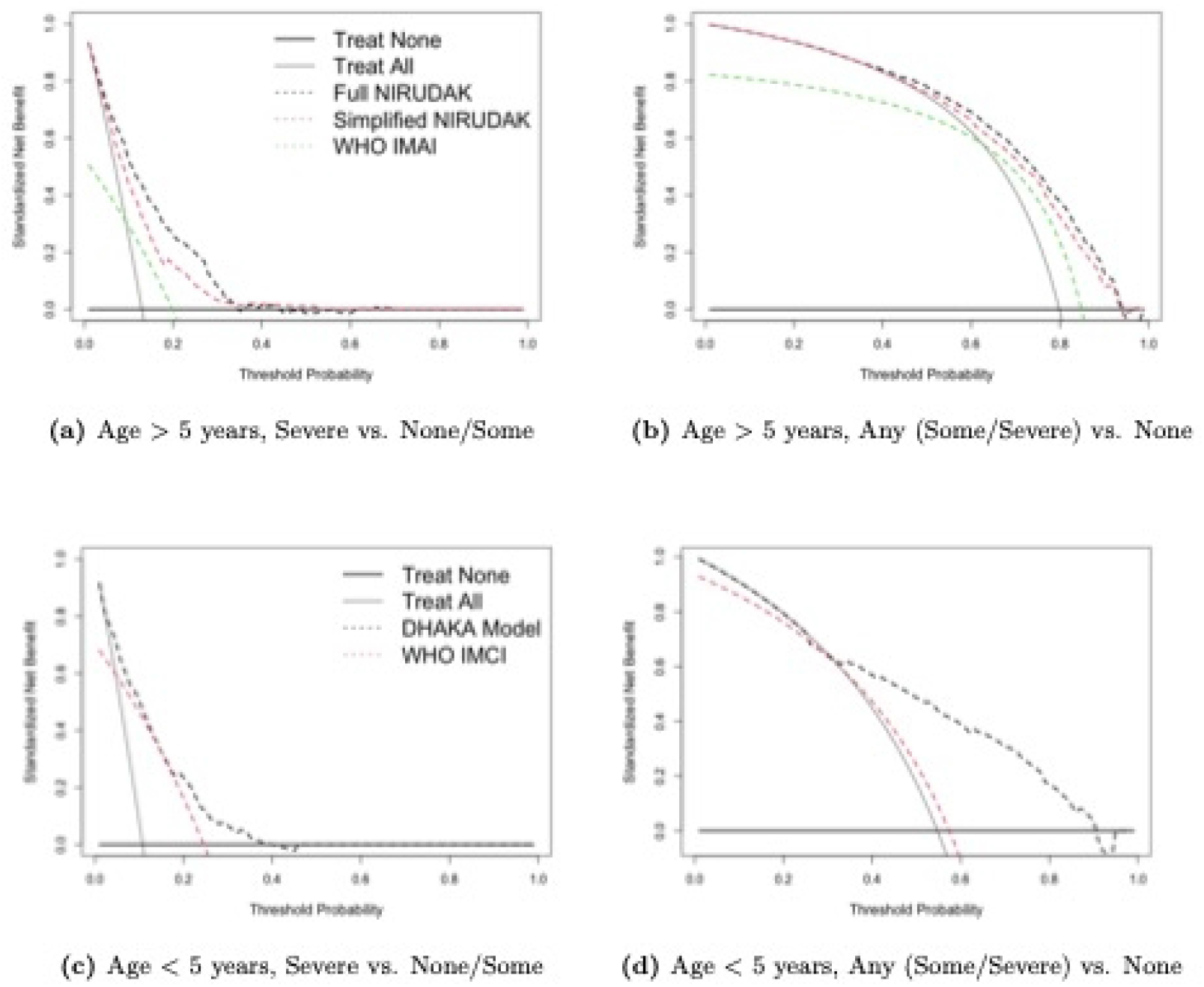
Standardized Net Benefit DCA for Dehydration models for individuals age *>* 5 years (NIRUDAK models & WHO IMAI algorithm) and age *<* 5 years (DHAKA model & WHO IMCI algorithm).

The decision curve analysis shows then that the DHAKA and NIRUDAK models provide the greatest benefit in terms of correctly identifying individuals with severe and any dehydration (and thereby minimizing the harm due to misidentifying truly dehydrated individuals).

Because the DHAKA and NIRUDAK models dominate all the other models over the whole range of clinically relevant thresholds for each binary split of the ordinal outcome, it is not necessary to compute areas under the curves to rank them as is necessary when one model does not dominate. However, one might still be interested in how much better they are over a range of thresholds that different decision makers or a single decision-maker might choose. Once one wants to draw conclusions more generally to a set of clinicians or other decision makers it is necessary to account for variation in their cost-benefit preferences. Determining this requires integrating net benefit over the relevant clinical threshold range with respect to the threshold distribution *f* (*r*) to obtain the weighted area under the net benefit curve.

This distribution of clinically relevant thresholds reflects how different decision makers value relative costs and benefits of treatment. Determining this distribution entails obtaining information on clinician/patient preferences or cost of treatment external to the study outcomes and risk predictions needed for model development. It therefore limits one of the primary advantages originally advocated for an individual decision maker’s use of the decision curve method [13]– namely, that it required only points on the decision curves corresponding to one’s own preferences.

For the purposes of this analysis, we adopt an approach more akin to a sensitivity analysis. That is, rather than empirically deriving and estimating *f* (*r*), we examine how the areas under the curve vary assuming different risk threshold distributions.

First, we assume *f* (*r*) is uniformly distributed over the risk threshold range of interest. Thus, *r* ∼ *U* (0.05, 0.15) and *r* ∼ *U* (0.6, 0.8) for severe and any dehydration, respectively. These distributions represent scenarios where a clinician might agree to treat a patient at any of the risk thresholds in each respective range with equal probability.

Second, we assume *r* ∼ *N* (0.10, 0.025) and *r* ∼ *N* (0.70, 0.05) for severe and any dehydration, respectively, where *N* (*a, b*) indicates a normal distribution with mean *a* and standard deviation *b*. These distributions, labeled Normal_1_, are centered at and give greatest weight to the midpoint of the clinically-reasonable ranges and assume 95% of clinically reasonable thresholds are between the bounds of the uniform threshold distributions with continuously decreasing weights as thresholds move away from the midpoint.

Next, we assume *r* ∼ *N* (0.07, 0.01) and *r* ∼ *N* (0.65, 0.01) for severe and any dehydration, respectively, and label these distributions Normal_2_. Normal_2_ represents a scenario where clinicians have lower average thresholds that do not have much variation, i.e., they are more risk averse compared to clinicians following the Normal_1_ distribution.

Finally, we examine a scenario in which clinicians have higher treatment thresholds. Scenario Normal_3_ assumes that *r* ∼ *N* (0.12, 0.02) and *r* ∼ *N* (0.75, 0.02) for severe and any dehydration, respectively.

The integrated weighted area also requires the outcome importance weights *w*. Because the consequences of missing cases of severe dehydration are more serious than the consequences of incorrectly diagnosing dehydration when it is not present, the total NB for diagnosing severe dehydration should be weighted more than that for diagnosing any dehydration. For example, we might weight the NB using the binary model for severe dehydration twice as much as that for any dehydration. A Beta(20,10) distribution for the relative weights has a mean of 2*/*3 with 95% of the distribution lying between about 0.5 and 0.8. This distribution assumes that most clinicians value a correct decision for diagnosing severe dehydration between 1 and 4 times that for diagnosing any dehydration (or equivalently no dehydration). Equivalently, it represents a single clinician’s uncertainty about these weights. This seems to be a reasonable first approximation.

Tables 4 and 5 give the Weighted Area Under the sNB Curves, *wAUC*_*sNB*_, using the uniform and three normal risk threshold distributions applied to the DHAKA, NIRUDAK and WHO models, as well as the Treat All and Treat None strategies for the two binary outcome comparisons. They also show the Integrated *wAUC*_*sNB*_ summary measure derived with the importance weights generated from a Beta(20,10) distribution.

**Table 4.** Weighted Area Under the sNB Curve for severe and any dehydration outcomes (95% CI) and Integrated Weighted Area Under sNB Curves (for *w*_*Severe*_ ∼*Beta*(20, 10) for the DHAKA model and WHO IMCI algorithm by risk threshold distribution, *f* (*r*).

| f(r)/Model | $wAUC_{sNB}$ | | Integrated $wAUC_{sNB}$ |
| --- | --- | --- | --- |
|  | Severe vs. (None/Some) | Any vs. None |  |
| **Uniform** | **$r \sim U(0.05, 0.15)$** | **$r \sim U(0.60, 0.80)$** |  |
| DHAKA Model | 0.50 (0.40, 0.60) | 0.30 (0.23, 0.37) | 0.43 (0.35, 0.51) |
| WHO IMCI | 0.46 (0.34, 0.58) | -0.77 (-1.02, -0.51) | 0.05 (-0.07, 0.17) |
| Treat All | 0.07 (-0.14, 0.30) | -1.03 (-1.31, -0.74) | -0.30 (-0.49, -0.09) |
| Treat None | 0 | 0 | 0 |
| **Normal<sub>1</sub>** | **$r \sim N(0.10, 0.025)$** | **$r \sim N(0.70, 0.05)$** |  |
| DHAKA Model | 0.50 (0.39, 0.61) | 0.31 (0.23, 0.38) | 0.43 (0.35, 0.52) |
| WHO IMCI | 0.46 (0.35, 0.58) | -0.74 (-0.98, -0.48) | 0.06 (-0.05, 0.18) |
| Treat All | 0.07 (-0.14, 0.30) | -0.99 (-1.26, -0.70) | -0.28 (-0.47, -0.07) |
| Treat None | 0 | 0 | 0 |
| **Normal<sub>2</sub>** | **$r \sim N(0.07, 0.01)$** | **$r \sim N(0.65, 0.01)$** |  |
| DHAKA Model | 0.60 (0.50, 0.69) | 0.36 (0.28, 0.43) | 0.52 (0.44, 0.59) |
| WHO IMCI | 0.54 (0.43, 0.65) | -0.36 (-0.55, -0.16) | 0.24 (0.14, 0.34) |
| Treat All | 0.37 (0.23, 0.52) | -0.54 (-0.75, -0.32) | 0.07 (-0.07, 0.21) |
| Treat None | 0 | 0 | 0 |
| **Normal<sub>3</sub>** | **$r \sim N(0.12, 0.02)$** | **$r \sim N(0.75, 0.02)$** |  |
| DHAKA Model | 0.44 (0.33, 0.55) | 0.25 (0.17, 0.33) | 0.38 (0.29, 0.46) |
| WHO IMCI | 0.42 (0.29, 0.54) | -1.16 (-1.47, -0.84) | -0.11 (-0.25, 0.03) |
| Treat All | -0.11 (-0.36, 0.16) | -1.50 (-1.85, -1.14) | -0.57 (-0.80, -0.32) |
| Treat None | 0 | 0 | 0 |

**Table 5.**
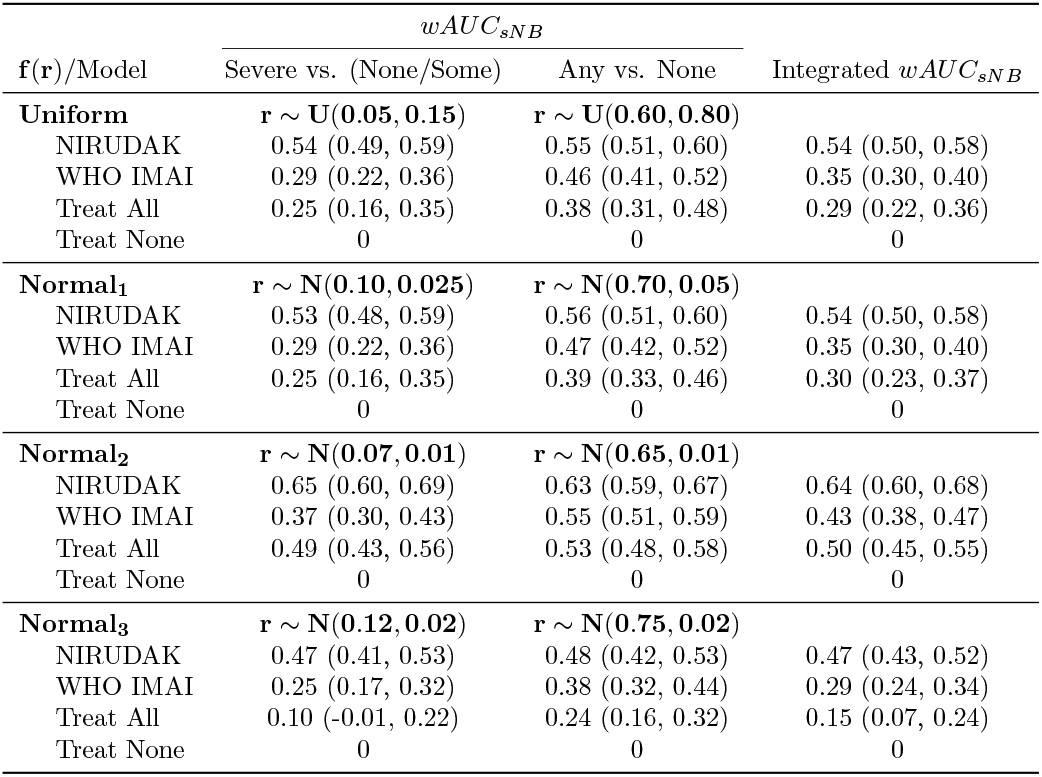
Weighted Area Under the sNB Curve for severe and any dehydration outcomes (95% CI) and Integrated Weighted Area Under sNB Curves (for *w*_*Severe*_ ∼ *Beta*(20, 10)) for NIRUDAK models and WHO IMAI algorithm by risk threshold distribution, *f* (*r*).

We see from Table 4 that the DHAKA model possesses the best *wAUC*_*sNB*_ for both binary comparisons, as well as the best Integrated *wAUC*_*sNB*_ for each assumed risk threshold distribution among children under five years of age. The Integrated NB ranges from 38% to 52% of the maximum possible achievable benefit across the four risk threshold distributions examined.

Table 5 shows that the NIRUDAK model is best for people five years of age and older for both binary comparisons and also achieves the greatest Integrated NB ranging from 47% to 64% of the maximum possible achievable NB for different *f* (*r*).

To assess the dependence of the final decision on the choice of importance weights, it is also helpful to plot the curve representing the total sNB for a range of potential importance weights that a single clinician might give. Fig 4 and Fig 5 display the Integrated *wAUC*_*sNB*_ for the WHO, DHAKA and NIRUDAK models for single importance weights varying between zero and one corresponding to the weight *w*_*severe*_ for the outcome of severe dehydration compared to that of any dehydration. The black triangles denote the total sNB when the two outcomes are equally weighted. Again, in each scenario the DHAKA and NIRUDAK models are better than the WHO models no matter which importance weights are used.

**Fig 4.**
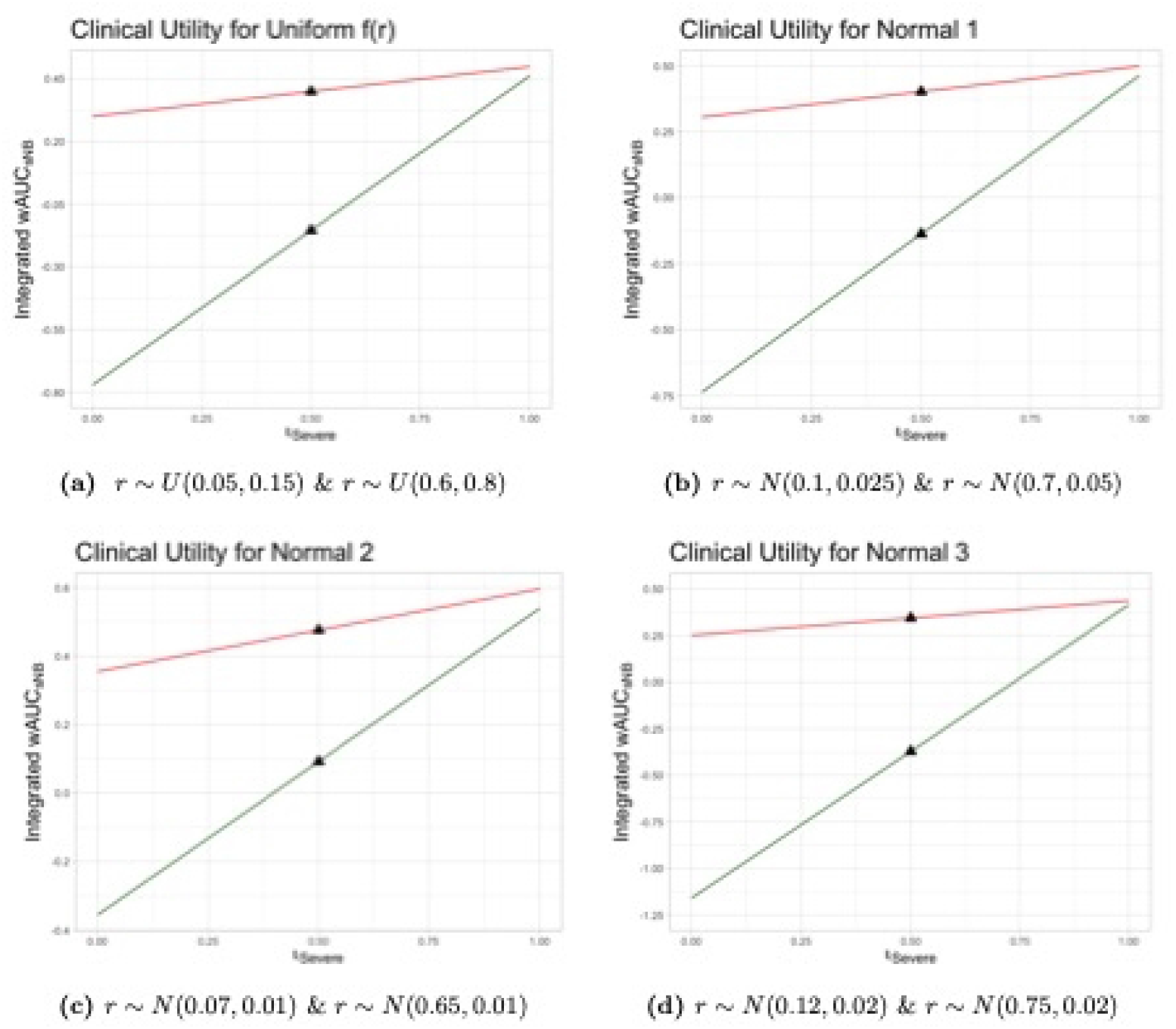
Integrated Weighted Area Under sNB Curves of DHAKA model and WHO IMCI by risk threshold distribution, *f* (*r*) (*r*_*Severe*_ *& r*_*Any*_), and importance weights for severe dehydration, *w*_*severe*_. DHAKA model (red) and WHO IMCI (green).

**Fig 5.**
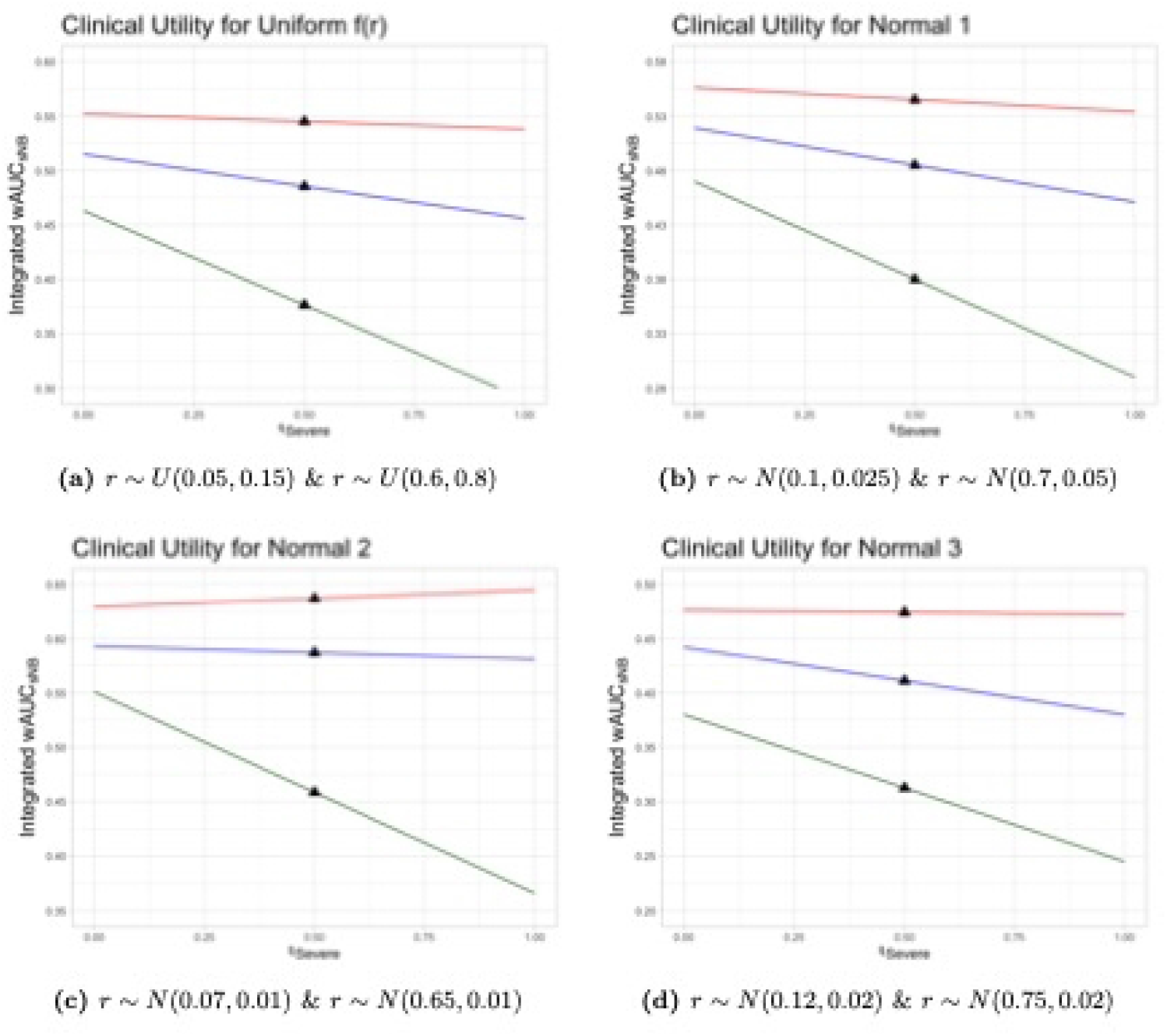
Integrated Weighted Area Under sNB Curves of NIRUDAK models and WHO IMAI by risk threshold distribution, *f* (*r*) (*r*_*Severe*_ *& r*_*Any*_), and importance weights for severe dehydration, *w*_*severe*_. NIRUDAK (red) and WHO IMAI (green).

## Discussion

In this paper, we have illustrated DCA using three ordinal logistic regression models, each of which estimated the probability of increasing levels of dehydration severity (none, some, severe) among individuals in Bangladesh presenting to a health clinic with acute diarrhea. Each model produced two binary decision curves formed by collapsing the ordinal outcome into the two binary comparisons of severe versus some or no dehydration, and any (severe or some) versus no dehydration. In order to obtain a measure of clinical utility for a model with a polytomous outcome, we developed and subsequently applied the weighted area under the standardized net benefit curve to each binary dehydration comparison and computed an average weighted by clinical importance to obtain an overall summary measure for each model, the Integrated Weighted Area Under sNB Curves.

The NIRUDAK model had greater NB across the range of reasonable risk threshold probabilities than the WHO IMAI algorithm for both binary comparisons among patients over five years of age. The DHAKA model also had greater clinical utility than the WHO IMCI algorithm for both dehydration comparisons across the appropriate range of risk thresholds for patients less than five years of age.

Though the same model was best for both age cohorts for both binary groupings, one could imagine a model that performs poorly in one dichotomization and relatively better in another. Therefore, we described a method for incorporating importance weights that reflect the relative clinical importance of the decisions to be based on different binary groupings of the outcome levels.

Since both the weighted area under the standardized net benefit curve, *wAUC*_*sNB*_, and the integrated weighted area, *IwAUC*_*sNB*_, can be viewed as an average sNB, they have the same units as sNB. When their values fall between zero and one, they can be interpreted as the proportion of maximum possible net benefit achieved. For instance, assuming a uniform distribution of *r*, the model for severe vs. not severe dehydration for the NIRUDAK data achieves 54% of the maximum possible achievable NB. Moreover, assuming a Beta(20,10) distribution for the weighting of the importance of the outcome for severe disease relative to the outcome of any disease leads to an integrated weighted standardized net benefit of 54% for the cumulative logit NIRUDAK model.

On the other hand, the negative net benefit for the WHO IMCI algorithm and Treat All strategies for the any dehydration outcome in DHAKA seen in Table 4 indicate that these two strategies have lower net benefit than Treat None, yielding harm rather than benefit. The negative values indicate that these treatment strategies lead to harm rather than benefit and so should be avoided.

In fact, both the NIRUDAK and DHAKA models were more clinically beneficial for any choice of importance weights. These models also had better calibration and discrimination than the WHO algorithms [6, 7, 20].

Both *wAUC*_*sNB*_ and *IwAUC*_*sNB*_ require estimating the distributions of predictive probability thresholds and importance weights that different clinicians consider relevant for making decisions. These may be challenging to determine in practice. For example, Talluri and Shete [14] proposed a method for determining threshold distributions that requires knowledge of the clinical decisions made using a specific prediction model in an external study that may not be relevant to the current situation. The potential subjectivity introduced by the choice of these distributions may introduce considerable uncertainty into the final analysis. It might be much easier for single decision-makers to describe their own uncertainty because it could be done internally.

The method we have proposed relies on dichotomizing a polytomous outcome into several binary sets and evaluating the utility on each set separately prior to averaging each result. Other approaches may be worth exploring, however. In the context of discrimination, for example, efforts have been made to transition from assessing pairwise comparisons of these binary sets (e.g., generalized *c*-index) to assessing sets of cases from each category simultaneously (e.g., ordinal volume under the ROC surface (VUS), ordinal *c*-index) [21]. The latter approach takes into account the full complexity of the response variable type, whereas the former examines the outcome in overlapping subsets. A similar multivariate extension of net benefit to a surface may also be informative.

Despite these limitations, our new measure has its strengths, as well. First, a single summary measure for clinical utility allows for straightforward comparison of the utility of two or more prediction models with polytomous outcomes. Second, the method naturally builds upon the robust methodology of net benefit and decision curve analysis and, similarly, can be applied to the data on which the model was derived or validated without the need for additional information, such as individual patient preferences.

Furthermore, the *IwAUC*_*sNB*_ has a simple and meaningful interpretation as the average net increase in the proportion of appropriately treated patients, weighted by the relative importance of each outcome of interest. The new measure also incorporates features that facilitate using the model in practice, from defining the appropriate risk threshold distribution to defining importance weights. The method allows for considerable flexibility to examine robustness of the findings to different model and feature choices that mimic clinical scenarios.

Lastly, an essential strength of this new extension is that it works for both ordered and unordered outcomes, as well as any polytomous prediction model. While we have used the the cumulative logit model for dehydration severity in our example, choosing a different model, for example choosing a different link function or removing the proportional odds assumption, will only change the predicted probabilities. While changing the model form might then lead to a different choice of the model that has the greatest overall utility, the method itself remains the same. Our approach is both flexible and robust and, thus, can be straightforwardly applied in practice.

## Conclusion

We developed a summary measure for evaluating the clinical utility of multi-category prediction models that was able to distinguish two predictive models as being superior to an existing standard for the management of dehydration in patients with acute diarrhea. This method could potentially be used to evaluate the clinical utility for other clinical prediction models with polytomous outcomes that are currently in common use by clinical providers worldwide.

## Data Availability

The de-identified NIRUDAK Study data set used for this secondary analysis is available on Open Science Framework. Link: https://osf.io/pncms/.

## Abbreviations

AUC_NB_: Area Under the NB Curve
DCA: Decision Curve Analysis
DHAKA: Dehydration Assessing Kids Accurately
FP: false-positives
HUM: hypervolume under the manifold
icddr,b: International Centre for Diarrhoeal Disease Research, Bangladesh
IMAI: Integrated Management of Adolescent and Adult Illness
IMCI: Integrated Management of Childhood Illness
IwAUC_sNB_: Integrated Weighted Area Under the Standardized NB Curve
NB: Net Benefit
NIRUDAK: Novel, Innovative Research for Understanding Dehydration in Adults and Kids
OSA: obstructive sleep apnea
OVO: One-Vs-One
OVR: One-Vs-Rest
ROC: receiver operating characteristic
SVR: Some Versus Rest
SNB: Standardized NB
TP: true-positives
VUS: volume under the ROC surface
wAUC_sNB_: Weighted Area Under the Standardized NB Curve
WHO: World Health Organization

## Acknowledgments

The authors would like to thank all study participants and study staff at icddr,b’s Dhaka Hospital for their help and support.

## Ethics approval and consent to participate

Ethical approval for both the DHAKA Study and the NIRUDAK study were obtained from the icddr,b’s Research Review Committee and Ethical Review Committee and the Rhode Island Hospital’s Institutional Review Board. All methods were performed in accordance with relevant guidelines and regulations. Formal written consent was obtained from each participant and/or their parent/guardian if under 18 years old in their native language, Bangla.

## Authors’ contributions

M.G., S.N., M.M., N.H.A. contributed to the study’s investigation. M.G., S.N., and M.M. contributed to data curation. A.N.Q., A.C.L, and C.H.S. contributed to the study’s methodology and conducted the formal analysis. A.N.Q. wrote the code for analysis. A.N.Q. and C.H.S. wrote the original draft. A.C.L. and C.H.S. contributed to the study’s conceptualization and supervision. A.C.L. contributed to funding acquisition. All authors reviewed and edited the manuscript.

